# Accessing Medical Care for Infertility: A Study of Women in Mexico

**DOI:** 10.1101/2022.02.02.22270322

**Authors:** Leslie V. Farland, Sana Khan, Stacey A. Missmer, Dalia Stern, Ruy Lopez-Ridaura, Jorge E. Chavarro, Andres Catzin-Kuhlmann, Ana Paola Sanchez-Serrano, Megan S. Rice, Martín Lajous

## Abstract

**Background:** Investigating the burden of access to infertility treatment has primarily been conducted in high-income countries, with little known for low- and middle-income countries, which comprise 80% of the world’s population. The objective of this study was to investigate access to infertility care in Mexico.

**Methods:** This was a cross-sectional analysis in the Mexican Teachers’ Cohort (MTC), a prospective cohort study of 115,307 Mexican female public school teachers from 12 states in Mexico. Log-binomial models, adjusted for age, hormonal contraceptive use, teaching in a rural school, and speaking an indigenous language, were used to estimate the prevalence ratio (PR) and 95% confidence intervals (95% CI) of accessing medical care for infertility among women reporting a history of infertility.

**Results:** 19,580 (17%) participants reported a history of infertility. Of those who experienced infertility, 12,470 (63.7%) reported seeking medical care for infertility, among whom 8,467 (67.9%) reported undergoing fertility treatments. Among women who reported a history of infertility, women who taught in a rural school (PR:0.95;0.92-0.97), spoke an indigenous language (PR:0.88; 0.84-0.92), or had less than a university degree (PR:0.93; 0.90-0.97) were less likely to access medical care for fertility. Women who had ever had a mammogram (PR:1.07; 1.05-1.10), had a pap-smear in the past year (PR:1.08;1.06-1.10), or who had utilized private healthcare regularly or in times of illness were more likely to access medical care for fertility.

**Conclusion:** Utilization of infertility care varied by demographic and access characteristics, including speaking an indigenous language, teaching in a rural school, and having a private healthcare provider.

## Background

Infertility affects approximately 50-80 million people worldwide; however, the true global burden is difficult to estimate given differing definitions of infertility and a lack of available surveillance data in many settings [1-3]. Among couples who experience infertility, there are many barriers that prevent them from accessing appropriate fertility care. Differences in access have been documented by race, age, socioeconomic status, and health-related factors [4-13]. Prior research on barriers to accessing fertility care has focused predominantly on the influence of markers of financial access (e.g. household income, insurance, education) and racial disparities in accessing care, with little information on how other cultural or lifestyle factors (e.g. physical activity, health history) may influence accessing fertility care [4, 14]. Moreover, the vast majority of the research on barriers to accessing fertility care has been conducted within the United States.

Low- and middle-income countries make up over 80% of the world’s population, but very little is known regarding the burden of infertility and access to fertility care in these settings. Extrapolating information from the U.S. to inform health care interventions in other regions is inappropriate due to differences in cultural and regional barriers to access. Prior research has suggested regional and geographic variations of infertility prevalence among couples in Mexico, possibly influenced by differences in economic factors, environmental exposures, literacy, and nutrition [15]. Therefore, the objective of this study was to investigate predictors of access to fertility care among a large cohort of reproductive-aged women across 12 states in Mexico enrolled in the Mexican Teacher’s Cohort (n=115,307) [16].

## Methods

### Study Design

The Mexican Teachers Cohort (MTC) is a large, prospective cohort study that was established in 2006 when teachers from two states (Veracruz and Jalisco) responded to a baseline questionnaire about their health and lifestyle [16]. The cohort was expanded to 10 additional states from 2008-2010 and includes 115,307 female teachers from across 12 diverse states in Mexico. The cohort study was a result of a partnership with Mexico’s public education system and included a range of culturally and economically diverse women. The MTC collected comprehensive baseline data of medical and lifestyle factors and assessed several exposures and risk factors associated with chronic disease. All participants have healthcare coverage, which includes fertility services, by a small number of social security institutions with integrated or separate healthcare providers. The study was approved by the institutional review board at the National Institute of Public Health in Mexico, and informed consent was obtained from all women. Our sample was restricted to participants who indicated that they had ever experienced infertility (n=19,580), and therefore, participants in the MTC who had not experienced infertility were excluded.

### Infertility History

Women were asked if they had ever undergone 12 months of trying to conceive without success (infertility). If they answered “yes” they were asked if they ever sought medical care for help to get pregnant and at what age they experienced infertility. They were then asked about what were the medical reason(s) why they experienced difficulty getting pregnant and were given the following possible responses: blocked tubes, polycystic ovary syndrome (PCOS), other ovulation disorders, endometriosis, abnormalities of the uterus, problems with the male partner, no known reason, and other. Participants could mark multiple reasons for their infertility. Participants with a history of infertility were then asked whether they received a medical treatment or procedure for help getting pregnant. Participants could select multiple treatment options including: none, intrauterine insemination (IUI), in vitro fertilization (IVF), and medications to induce ovulation (clomiphene, metformin, injections of gonadotropin, other treatment or procedure). For this analysis, participants were categorized as having “accessed medical care for infertility” if they reported that they sought medical attention for themselves or their partner to achieve pregnancy or if they reported having utilized fertility treatment or had a diagnosis for their infertility.

### Demographic predictors of accessing infertility care

Information on demographic characteristics was assessed on the baseline questionnaire in 2008. Specifically, we collected information on teaching in a rural school (no, yes), speaking an indigenous language (no, yes), highest level of education completed (less than university, university degree, graduate degree), and state of residence which we categorized into four regions (Mexico City, northern states (Baja California, Durango, Nuevo León, Sonora), central states (Guanajuato, Hidalgo, Jalisco, México), and southern states (Chiapas, Yucatán, Veracruz)).

### Health systems predictors of accessing infertility care

We also collected information on markers of health care utilization. In Mexico, federal and state-level employees and individuals in the formal private sector have healthcare coverage through several social security systems. ISSSTE (Instituto de Seguridad y Servicios Sociales de los Trabajadores del Estado) covers federal government employees (79.5% of MTC participants), IMSS (Instituto Mexicano del Seguro Social) is responsible for the care of the majority of state-level employees in the cohort (11.4%), while four more public healthcare providers do the same for the remaining state-level employees (9.1%). However, a participant covered by one social security institution may choose to seek care with a private provider or with a different social security institution, that provides care to a family member. Thus, independently of women’s employer and social security coverage, we categorized women according to their self-reported health services used for regular care (private, IMSS, ISSSTE, other public, other) and health service used for major illness or intervention (private, IMSS, ISSSTE, other public, other). We also collected information on history of mammogram (never, ever) and pap-smear in the past year (no, yes) which we considered proxy variables for access to healthcare and screening services.

### Reproductive and lifestyle predictors of accessing fertility care

Finally, we collected information on health and lifestyle history at baseline including age at menarche (< 12, 12, 13, ≥ 14), hormonal contraceptive usage (never, ever), parity (nulliparous, 1, 2, 3+), history of smoking (never, ever), alcoholic drinks/day (0, <0.1, ≥ 0.1), participated in vigorous physical activity at age 18 (≤ 3 hours per week, > 3 hours per week), and history of type 2 diabetes (no, yes), given its association with polycystic ovary syndrome (PCOS). Hormonal contraceptive use was investigated as covariate in our analysis because it reflects having a connection with the medical system and is suggestive of pregnancy planning. Information on height and weight at baseline and at age 18 was used to calculate body mass index (BMI; kg/m^2^) (BMI in 2008: <25, 25-<30, ≥ 30) (BMI at age 18: <18.5, 18.5-<21, 21-<25, ≥ 25). Body size was estimated based on figure drawings (somatotypes) [17]. Women were asked to report the figure drawing (range 1-9) that best reflected their body shape in young adolescence (2 years after their first menstrual period) and when they were 25-30 years old. For our analysis, we created categories of somatotype in young adolescence (1, 2, 3, ≥4) and somatotype at 25-30 years (1-3, 4, 5, ≥6).

### Statistical Analysis

For the analysis, we utilized data collected at study baseline in 2008. Among women with a history of infertility, we modeled the probability of accessing medical care for infertility. Generalized linear models with a log link and a binomial distribution were used to estimate the prevalence ratio (PR) and 95% confidence intervals (CI) of seeking medical care for infertility. Multivariable models were adjusted for age, hormonal contraceptives (HC) use, teaching in rural school, and speaking an indigenous language. These covariates were chosen for adjustment in multivariable models given their strong observed relationship with accessing fertility care in crude models. For covariates with missing values, missing indicators variables were created. SAS version 9.4 (Carry, NC, USA) was used to conduct these analyses.

## Results

Among the 115,307 participants, 19,580 (17%) reported infertility. Participants who reported having accessed medical care for infertility were, on average, 43.2 years old (SD=7.0) at baseline and 28.0 (5.3) at first experience of infertility, while participants who did not access care for infertility were 44.2 (7.3) years old at baseline and 26.1 (5.6) at reported infertility. Among women who experienced infertility, 63.7% (n=12,470) reported accessing medical care for infertility (Table 1). Among women who did access care, the most common infertility diagnoses were ovulatory disorders (other than PCOS) (18.7%), tubal-factor infertility (16.2%), and PCOS (13.0%); 21.5% reported an unknown or idiopathic cause of their infertility and 11% reported cause attributed to their male partner. The majority of participants with infertility reported having used fertility treatment (67.9%). The most common type of treatment utilized was ovulation induction (62.3%), with fewer women reporting IUI (4.3%) and IVF (1.3%). When asked about specific fertility drugs used, the majority of women reported using “other” (47.5%), followed by clomiphene (34.5%), and gonadotropin injections (14.6%).

**Table 1.** Accessing medical care for infertility among participants with self-reported infertility in the Mexican’s Teacher’s Cohort at baseline in 2008

|  |  | Accessed medical care for infertility |  |
| --- | --- | --- | --- |
|  |  | No<br>(n=7,110) | Yes<br>(n=12,470) |
| Infertility experience |  |  |  |
| Age at reported infertility* |  | 26.1(5.6) | 28.0(5.3) |
| Type of infertility diagnosis |  |  |  |
|  | Fallopian tube, % |  | 16.2 |
|  | Polycystic ovarian syndrome, % |  | 13.0 |
|  | Ovulatory infertility, % |  | 18.7 |
|  | Endometriosis, % |  | 7.9 |
|  | Uterine factor infertility, % |  | 7.2 |
|  | Male factor infertility, % |  | 11.0 |
|  | Unknown infertility, % |  | 21.5 |
|  | Other reason, % |  | 13.4 |
| Type of infertility treatment used |  |  |  |
|  | - None, % |  | 32.1 |
|  | - Intrauterine Insemination (IUI), % |  | 4.3 |
|  | - In Vitro Fertilization (IVF), % |  | 1.3 |
|  | - Ovulation Induction, % |  | 62.3 |
| Among those who utilized treatment, type of infertility drugs used |  |  |  |
|  | - Clomiphene, % |  | 34.5 |
|  | - Metformin, % |  | 3.4 |
|  | - Gonadotropin Injections, % |  | 14.6 |
|  | - Other, % |  | 47.5 |
| Demographic characteristics |  |  |  |
| Age (yrs)* |  | 44.2(7.3) | 43.2(7.0) |
| Body Mass Index (BMI)(kg/m2) |  | 27.8(4.5) | 27.9(4.6) |
| Nulliparous, % |  | 8.6 | 19.3 |
| Smoking history |  |  |  |
|  | - No, % | 76.7 | 76.5 |
|  | - Current/former, % | 23.3 | 23.5 |
| History of hormonal contraceptive use, % |  | 46.2 | 39.1 |
| Age at first birth* |  | 25.4(4.5) | 26.8(4.7) |
| Vigorous activity >3 hours at age 18, % |  | 74.0 | 76.0 |
| Somatotype Adolescent (2 years after your first menstrual period) |  |  |  |
|  | - 1, % | 14.1 | 14.7 |
|  | - 2, % | 27.2 | 29.6 |
|  | - 3, % | 28.3 | 27.7 |
|  | - 4, % | 23.0 | 23.6 |
|  | - 5, % | 7.5 | 4.4 |
| BMI at age 18 |  |  |  |
|  | - <18.5, % | 18.7 | 18.7 |
|  | - 18.5-<21, % | 34.5 | 34.3 |

|  | Accessed medical care for infertility |  |
| --- | --- | --- |
|  | No<br>(n=7,110) | Yes<br>(n=12,470) |
| - 21->25, % | 37.1 | 37.0 |
| - ≥25, % | 9.7 | 10.1 |
| Age at menarche |  |  |
| - ≤12, % | 24.4 | 26.2 |
| - 12, % | 27.9 | 29.0 |
| - 13, % | 19.7 | 19.9 |
| - ≥14, % | 28.0 | 25.0 |
| Health service used (Regular care) |  |  |
| - Private, % | 17.4 | 23.5 |
| - ISSSTE, % | 62.4 | 56.4 |
| - IMSS, % | 9.3 | 8.5 |
| - Other Public, % | 1.0 | 0.7 |
| - Other, % | 9.8 | 10.9 |
| Health service used (Illness) |  |  |
| - Private, % | 16.6 | 18.9 |
| - ISSSTE, % | 63.1 | 60.1 |
| - IMSS, % | 10.8 | 10.8 |
| - Other Public, % | 1.0 | 1.0 |
| - Other, % | 8.4 | 9.2 |
| Speaks and indigenous language, % | 10.7 | 7.6 |
| Teaches in a rural school, % | 26.0 | 22.6 |

Values are means(SD) or percentages and are standardized to the age distribution of the study population.
Values of polytomous variables may not sum to 100% due to rounding
\* Value is not age adjusted

When investigating the relationship between demographic characteristics and probability of fertility care several associations emerged (Table 2). Women who taught in a rural school (PR:0.95; 0.92-0.97), or who spoke an indigenous language (PR:0.88; 0.84-0.92) were less likely to access fertility care. Compared to women with a university degree, women with a graduate degree were more likely to access medical care for infertility (PR: 1.06; 1.03-1.09), while women with less than a university degree were less likely to access care (PR:0.93; 0.90-0.97). Compared to women who lived in Mexico City, women who lived in central states (Guanajuato, Hidalgo, Jalisco, and México) were also less likely to access fertility care (RR: 0.96; 0.93-0.99).

**Table 2.** The association between demographic characteristics and accessing medical care for infertility in the Mexican teacher’s cohort, among women reporting a history of infertility

|  | **Did not access**<br>**care for**<br>**infertility**<br>**7,110 (36.3%)** | **Accessed**<br>**medical care**<br>**for infertility**<br>**12,470 (63.7%)** | **Prevalence Ratio**<br>**(95% CI)**<br>**accessing care**<br>**for infertility** | **Multivariable**<br>**Adjusted**<br>**Prevalence Ratio<sup>1</sup>**<br>**(95% CI)** |
| --- | --- | --- | --- | --- |
| Teacher in a rural school |  |  |  |  |
| No | 5,222 (35.5) | 9,468 (64.5) | 1.00 (Ref) | 1.00 (Ref) |
| Yes | 1,783 (38.6) | 2,837 (61.4) | 0.95 (0.93-0.98) | 0.95 (0.92-0.97) |
| Speaks an indigenous<br>language |  |  |  |  |
| No | 6,263 (35.5) | 11,393 (64.5) | 1.00 (Ref) | 1.00 (Ref) |
| Yes | 793 (45.7) | 954 (56.4) | 0.87 (0.84-0.91) | 0.88 (0.84-0.92) |
| Highest level of<br>education completed |  |  |  |  |
| <University | 896 (40.8) | 1,300 (59.2) | 0.91 (0.88-0.94) | 0.93 (0.90-0.97) |
| University degree | 4,111 (35.0) | 7,648 (65.0) | 1.00 (Ref) | 1.00 (Ref) |
| Graduate degree | 709 (29.9) | 1,664 (70.1) | 1.08 (1.05-1.11) | 1.06 (1.03-1.09) |
| Region<sup>2</sup> |  |  |  |  |
| Mexico City | 1,023 (33.3) | 2,048 (66.7) | 1.00 (Ref) | 1.00 (Ref) |
| North | 1,309 (36.2) | 2,312 (63.9) | 0.96 (0.92-0.99) | 0.97 (0.94-1.01) |
| Central | 2,323 (37.9) | 3,805 (62.1) | 0.93 (0.90-0.96) | 0.96 (0.93-0.99) |
| South | 2,455 (36.3) | 4,305 (63.7) | 0.95 (0.93-0.98) | 0.98 (0.95-1.01) |

1) Multivariable models adjusted for age (continuous), history of hormonal contraceptive use, teaching in a rural school, speaking an indigenous language
2) North: Baja California, Durango, Nuevo León, Sonora;
Central: Guanajuato, Hidalgo, Jalisco, México;
South: Chiapas, Yucatán, Veracruz

When investigating the role of health systems access (Table 3), we found that women who had ever had a mammogram (PR:1.07; 1.05-1.10), or who had undergone a pap-smear in the past year (PR:1.08; 1.06-1.10) were more likely to access fertility care compared to women who had not (Table 4). Compared to women who utilized private health providers as their primary provider, women who utilized IMSS (PR: 0.88;0.86 -0.91) or ISSSTE (PR: 0.88; 0.84-0.92) as their primary health care provider were less likely to seek medical care for fertility, as were women with other public insurance (PR: 0.83; 0.72-0.95) and other insurance (PR: 0.94; 0.91-0.97). Specifically, during a time of illness, women who utilized IMSS (PR: 0.94; 0.92-0.97) or ISSSTE (PR: 0.95; 0.95-0.99) were less likely to seek out fertility care compared to women who utilized private health providers during times of illness.

**Table 3.** The association between health systems access and accessing medical diagnosis of infertility in the Mexican teacher’s cohort, among women reporting a history of infertility

|  | **Did not access**<br>**care for**<br>**infertility**<br>**7,110 (36.3%)** | **Accessed**<br>**medical care**<br>**for infertility**<br>**12,470 (63.7%)** | **Relative Risk**<br>**(95% CI)**<br>**accessing care**<br>**for infertility** | **Multivariable**<br>**Adjusted**<br>**Relative Risk<sup>1</sup>**<br>**(95% CI)** |
| --- | --- | --- | --- | --- |
| Health service used<br>(Regular care) |  |  |  |  |
| Private | 1,177 (29.5) | 2,811 (70.5) | 1.00 (Ref) | 1.00 (Ref) |
| IMSS | 4,262 (38.9) | 6,701 (61.1) | 0.87 (0.85-0.89) | 0.88 (0.86-0.91) |
| ISSSTE | 627 (38.3) | 1,011 (61.7) | 0.88 (0.84-0.91) | 0.88 (0.84-0.92) |
| Other Public | 69 (44.0) | 88 (56.1) | 0.80 (0.69-0.91) | 0.83 (0.72-0.95) |
| Other | 657 (33.5) | 1307 (66.6) | 0.94 (0.91-0.98) | 0.94 (0.91-0.97) |
| Health service used<br>(Illness) |  |  |  |  |
| Private | 1,123 (32.8) | 2,304 (67.2) | 1.00 (Ref) | 1.00 (Ref) |
| IMSS | 4,363 (37.6) | 7,228 (62.4) | 0.93 (0.90- 0.95) | 0.94 (0.92-0.97) |
| ISSSTE | 742 (36.1) | 1,311 (63.9) | 0.95 (0.91-0.99) | 0.95 (0.92-0.99) |
| Other Public | 67 (36.2) | 118 (63.8) | 0.95 (0.85-1.06) | 0.97 (0.87-1.09) |
| Other | 577 (34.3) | 1105 (65.7) | 0.98 (0.94-1.02) | 0.98 (0.94-1.02) |
| Mammogram |  |  |  |  |
| Never | 3,584 (37.0) | 6,096 (63.0) | 1.00 (Ref) | 1.00 (Ref) |
| Ever | 3,204 (34.8) | 6,015 (65.3) | 1.04 (1.01-1.06) | 1.07 (1.05-1.10) |
| Pap-smear in past year |  |  |  |  |
| No | 3,438 (38.9) | 5,391 (61.1) | 1.00 (Ref) | 1.00 (Ref) |
| Yes | 3,672 (34.2) | 7,079 (65.9) | 1.08 (1.06-1.10) | 1.08 (1.06-1.10) |

1) Multivariable models adjusted for age (continuous), history of hormonal contraceptive use, teaching in a rural school, speaking an indigenous language

**Table 4.** The association between reproductive and lifestyle characteristics and accessing medical care for infertility in the Mexican teacher’s cohort, among women reporting a history of infertility

|  | **Did not access**<br>**care for**<br>**infertility**<br>**7,110 (36.3%)** | **Accessed**<br>**medical care**<br>**for infertility**<br>**12,470 (63.7%)** | **Relative Risk**<br>**(95% CI)**<br>**accessing care**<br>**for infertility** | **Multivariable**<br>**Adjusted**<br>**Relative Risk<sup>1</sup>**<br>**(95% CI)** |
| --- | --- | --- | --- | --- |
| Hormonal contraceptive use |  |  |  |  |
| Never | 3,820 (33.5) | 7,597 (66.5) | 1.00 (Ref) | 1.00 (Ref) |
| Ever | 2,764 (39.6) | 4,210 (60.4) | 0.91 (0.89-0.93) | 0.91 (0.89-0.93) |
| Parity in 2008 |  |  |  |  |
| Nulliparous | 587 (19.9) | 2,358 (80.1) | 1.00 (Ref) | 1.00 (Ref) |
| 1 | 1,171 (32.7) | 2,413 (67.3) | 0.84 (0.82-0.87) | 0.85 (0.82-0.87) |
| 2 | 2,139 (38.6) | 3,396 (61.4) | 0.77 (0.75-0.79) | 0.78 (0.76-0.80) |
| 3+ | 3,062 (42.9) | 4,076 (57.1) | 0.71 (0.69-0.73) | 0.74 (0.72-0.76) |
| History of smoking |  |  |  |  |
| Never | 5,085 (35.6) | 9,185 (64.4) | 1.00 (Ref) | 1.00 (Ref) |
| Ever | 1,566 (36.0) | 2,779 (64.0) | 0.99 (0.97-1.02) | 1.00 (0.98-1.03) |
| Alcoholic drinks/day |  |  |  |  |
| 0 | 2,362 (36.1) | 4174 (63.9) | 1.00 (Ref) | 1.00 (Ref) |
| <0.1 | 2,780 (34.4) | 5,291 (65.6) | 1.03 (1.00-1.05) | 1.02 (1.00-1.05) |
| ≥0.1 | 1,111 (35.2) | 2,047 (64.8) | 1.01 (0.98-1.05) | 1.02 (0.99-1.05) |
| Type 2 Diabetes |  |  |  |  |
| No | 6,577 (36.2) | 11,593 (63.8) | 1.00 (Ref) | 1.00 (Ref) |
| Yes | 533 (37.8) | 877 (62.2) | 0.97 (0.93-1.02) | 1.00 (0.96-1.04) |
| Vigorous Physical Activity at age 18 |  |  |  |  |
| ≤ 3 hours per week | 1,422 (36.0) | 2,525 (64.0) | 1.00 (Ref) | 1.00 (Ref) |
| > 3 hours per week | 3,997 (33.2) | 8,026 (66.8) | 1.04 (1.02-1.07) | 1.04 (1.01-1.06) |
| Age at menarche |  |  |  |  |
| <12 | 1,690 (34.3) | 3,233 (65.7) | 1.00 (Ref) | 1.00 (Ref) |
| 12 | 1,931 (35.0) | 3,584 (65.0) | 0.99 (0.96-1.02) | 1.00 (0.97-1.03) |
| 13 | 1,391 (36.4) | 2,436 (63.7) | 0.97 (0.94-1.00) | 0.98 (0.95-1.01) |
| ≥14 | 1,993 (39.5) | 3,053 (60.5) | 0.92 (0.89-0.95) | 0.95 (0.92-0.98) |
| Linear Trend |  |  | <0.0001 | 0.0002 |
| BMI (kg/m<sup>2</sup>) in 2008 |  |  |  |  |
| <25 | 1,852 (35.1) | 3,429 (64.9) | 1.00 (Ref) | 1.00 (Ref) |
| 25-<30 | 2,781 (37.0) | 4,738 (63.0) | 0.97 (0.95-1.00) | 0.99 (0.96-1.01) |
| ≥30 | 1,753 (35.4) | 3,189 (64.5) | 0.99 (0.97-1.02) | 1.01 (0.98-1.04) |
| Linear Trend |  |  | 0.88 | 0.34 |
| Somatotype in young adolescence<sup>2</sup> |  |  |  |  |
| 1 | 1,002 (35.3) | 1,838 (64.7) | 1.00 (Ref) | 1.00 (Ref) |
| 2 | 1,936 (34.4) | 3,686 (65.6) | 1.01 (0.98-1.05) | 1.01 (0.98-1.05) |
| 3 | 2,006 (36.8) | 3,451 (63.2) | 0.98 (0.94-1.01) | 0.98 (0.95-1.01) |
| ≥4 | 1,626 (35.5) | 2,955 (64.5) | 1.00 (0.96-1.03) | 1.00 (0.96-1.03) |
| Linear Trend |  |  | 0.28 | 0.24 |
| BMI at age 18 |  |  |  |  |
| <18.5 | 1,099 (35.8) | 1,967 (64.2) | 1.00 (Ref) | 1.00 (Ref) |
| 18.5-<21 | 2,018 (35.8) | 3,615 (64.2) | 1.00 (0.97-1.03) | 1.00 (0.97-1.03) |
| 21- <25 | 2,164 (35.6) | 3,915 (64.4) | 1.00 (0.97-1.04) | 1.00 (0.97-1.04) |
| ≥ 25 | 556 (34.1) | 1,074 (65.9) | 1.03 (0.98-1.07) | 1.02 (0.97-1.06) |
| Linear Trend |  |  | 0.25 | 0.45 |
| Somatotype (25-30 y) |  |  |  |  |
| 1-3 | 1,659 (35.2) | 3,055 (64.8) | 1.00 (Ref) | 1.00 (Ref) |
| 4 | 1,714 (35.9) | 3,064 (64.1) | 0.99 (0.96-1.02) | 0.98 (0.96-1.01) |
| 5 | 1,132 (35.6) | 2,047 (64.4) | 0.99 (0.96-1.03) | 0.99 (0.95-1.02) |
| ≥6 | 2,060 (35.8) | 3,699 (64.2) | 0.99 (0.96-1.02) | 0.98 (0.95-1.01) |
| Linear Trend |  |  | 0.74 | 0.23 |

1) Multivariable models adjusted for age (continuous), history of hormonal contraceptive use, teaching in a rural school, speaking an indigenous language
2) Young adolescence defined as 2 years after first menstrual period

When investigating the role of reproductive and lifestyle characteristics, women who had a history of using HCs were less likely to access care (PR:0.91; 0.89-0.93) (Table 4). Compared to nulliparous women, women who were parous were also less likely to access fertility care, with women who had three or more children the least likely to access care (PR: 0.74; 0.72 – 0.76). We observed a statistically significant inverse linear relationship between increasing age at menarche and likelihood of accessing medical care for infertility (P<0.001 for linear trend). Women who reported >3 hours of vigorous physical activity per week at age 18 were more likely to access fertility care (PR:1.04; 1.01-1.06). There was no difference between women seeking care for infertility by history of smoking, alcohol consumption, type 2 diabetes diagnosis, BMI at age 18 or baseline, or body size in adolescence or adulthood.

## Discussion

Among our cohort of female teachers across 12 regions in Mexico, 17% of women reported infertility. Of these women, the majority (63.7%), reported seeking access to fertility care. The most common diagnoses of infertility were ovulatory, tubal factor, and unknown. Utilization of fertility care in Mexico varied by demographic, lifestyle, and access characteristics. Women were less likely to seek access to infertility care if they were single, used HCs, taught in a rural area, spoke an indigenous language, or had less than a university degree. Women were also less likely to access medical care for infertility if they had previously had a child. Conversely, women were more likely to seek medical care for infertility if they had ever had a mammogram or pap-smear in the past year, or if they had utilized private health providers.

Our study found that approximately 17% of women in our cohort reported a history of infertility. This finding is slightly higher than estimates in the United States that have ranged from 6.0% [18] to 15.5% [19]. Estimates of infertility prevalence across 25 population surveys from low, middle, and high income countries observed that infertility ranged from 3.5% - 16.7% with an overall median prevalence among less developed countries of 9% [20]. The majority of infertile women in our sample accessed medical care for their infertility (63.7%). Our estimate of fertility care access is similar to previous findings from 25 international studies which found that 56% of couples reported access to medical care for infertility globally (range: 42-76%), with slightly fewer couples seeking care in less developed countries (mean=51.2%; range 27-74%)[20]. This is similar to findings from the Nurses’ Health Study in the United States (65%)[4, 21], but greater than estimates from the National Survey for Family Growth in the United States (36%) [18]. This may reflect the fact that Mexican citizens who are government employees or in the formal private sector have access to universal health coverage and therefore, are more likely access to medical care compared to couples in the United States. Among women who accessed medical care for infertility, the most common diagnoses were ovarian infertility, PCOS, blockage of the fallopian tube, and unknown. Our findings are consistent with prior research on infertility in Mexico that suggested that the most common causes for infertility were asymptomatic infection and anovulation, possibly indicative of PCOS [15]. Only 11% of women indicated that their infertility is due to the male partner, which is lower than previous estimates among infertile couples in Mexico [22]. Consistent with findings in United States [4], the cause of infertility for many women is unknown.

Of those women who accessed medical care for fertility, the majority underwent ovulation induction (62.3%); the most common drug utilized was clomiphene (34.5%). A large percentage of women (47.5%) reported using “other” types of infertility drugs. This may reflect differences in drug name provided on the survey (e.g. Clomifeno) and the more commonly known brand names for clomiphene which was not included on the questionnaire (e.g. Omifin). Of all the women who received treatment, only 1.3% underwent IVF. In México, there are only three public hospitals with in vitro fertilization program: Instituto Nacional de Perinatología (México City), Centro Médico Nacional 20 de Noviembre (Mexico City), and Hospital Materno Perinatal Mónica Pretelini (State of México). Therefore, women may need to wait to access or pay higher costs with private clinics which may explain the low utilization of IVF.

Among women who experience infertility, not all are able to access medical care to treat their infertility. Issues related to accessing medical care for fertility are complex. In the United States, there are established differences in accessing fertility care by race, age, causes of infertility, and socioeconomic factors that influence who receives medical care [4, 8]. Indeed, research from the National Survey of Fertility Barriers found that Black and Hispanic women were less likely to receive infertility services compared to white women and that this relationship was driven, but not fully accounted for by, income, insurance status, and level of education [9]. Research from the National Survey of Family Growth found that among women who reported infertility, whether a woman sought out fertility treatment varied by income, insurance coverage, age, and parity [6]. In addition to socioeconomic factors, other demographic, lifestyle, and environmental factors may also explain potential differences between women who accessed care and those who do not, however, this relationship has not been adequately studied. Prior research in the Nurses’ Health Study II found that in addition to the traditional relationships, a pattern of “healthy lifestyle behavior” was associated with accessing infertility care. Women were less likely to seek medical care related to infertility if they were older, parous, current smokers, or had a higher BMI than their counterparts who did seek medical care [4]. Those who did seek fertility care were also more likely to take multivitamins, exercise, and have had a recent physical examination.

We observed that women who reported speaking an indigenous language were less likely to access medical care for infertility. Prior research has suggested that indigenous people in Mexico have a higher prevalence of health problems and lower rates of utilizing primary health care [23]. Additionally, we found that women with graduate-level education were more likely to access care, while women with less than a university degree were less likely to access fertility care. Our findings support other studies that have found higher levels of education was associated with increased access to fertility care [12]. This gradient demonstrates the role education plays in gaining financial resources that may help access care but may also be reflective of self-advocacy skills learned from gaining a higher education.

We found that reproductive history and some lifestyle factors were associated with access to infertility care. Women who reported ever using HCs, were parous, and experienced older age at first menstrual period were less likely to access fertility care. We found no association between type 2 diabetes history, BMI at age 18 and questionnaire baseline, or body size in adolescence and adulthood and accessing fertility care. Women who participated in three or more hours of vigorous physical activity at age 18 were more likely to access fertility care. This finding is consistent with other research from the Nurses’ Health Study II that found a pattern with “healthy lifestyle behaviors.” Specifically, they found that women who exercised regularly were more likely to access fertility care [4].

The majority of women in our popular reported using ISSSTE, which covers health care for federal government employees, for regular health care needs and for major illness or intervention; our findings demonstrated that those who were able to supplement their public or government insurance (ISSSTE, IMSS, Other Public) with private health coverage were more likely to access fertility care. We also found that women who taught in a rural area were less likely to access fertility care indicating that women who live and teach in rural areas may be presented with additional geographic barriers to seeking care. Geographic barriers to accessing fertility care has been documented in the United States as well [24], as quality fertility services tend to be clustered in urban regions, women who live further away from these centers need to travel a greater distance to access this care. We found that women who had a mammogram or pap-smear in the past year, were more likely to access infertility care, suggesting that women who are more connected with the medical system (i.e. undergoing screening services) may be more likely to access fertility care. When stratified by region, women in the north, central, and south regions were less likely to report seeking access to infertility care, as compared to women in Mexico City.

Our study, among a cohort of Mexican women, confirmed similar patterns of access as have been found in other populations; women who are older in age, single, had lower income, had lower education levels, or who taught in rural areas were less likely to access medical care for infertility, while women who had private health insurance, had undergone mammography, or had a pap-smear in the past year were more likely to receive medical care. We found some unique patterns related to accessing fertility care among women in Mexico; women who spoke an indigenous language and who lived in regions outside of Mexico City were less likely to access care. Our findings add to the existing body of literature, which can inform future policy recommendations, by examining how lifestyle and demographic factors influence who receives care and provide insight into how these factors are related to accessing care. Future research should continue to investigate policies focused on improving access to fertility care for women who speak an indigenous language or who live in rural areas.

A strength of our study was the use of the Mexican Teacher’s Cohort, a well-established, large cohort study with detailed information from across 12 states in Mexico [16]. However, there also are important limitations of our findings. Our analysis utilizes self-reported measures that may be prone to misclassification. However, we would expect that any misclassification would be non-differential and thus, attenuate our reported relationships. Additionally, given the cross-sectional nature of the baseline survey collection, there is the possibility of recall bias, however we would expect this bias to be minimal. Additionally, the findings of the study may not be generalizable to other populations, as this cohort was comprised of women employed as teachers. Thus, these findings may be most appropriately generalized to women with similar occupational and educational backgrounds within Mexico with access to healthcare. However, our population has geographic variability as we were able to study women from 12 states and a number of geographic regions across Mexico.

## Conclusion

In sum, we found that utilization of fertility care varied by demographic, lifestyle, and access characteristics, including speaking an indigenous language, teaching in a rural school, and having a private healthcare provider. These findings could inform public health policy on alleviating barriers in access to care among infertile women in Mexico.

## Data Availability

The data that support the findings of this study are available, but restrictions apply to the availability of these data, which were used under license for the current study, and so are not publicly available. Data are however available from the authors upon reasonable request and with permission.

## Declarations

### Ethical Approval and Consent to participate

The study was approved by the institutional review board at the National Institute of Public Health in Mexico, and informed consent was obtained from all women.

### Authors’ contributions

LVF, MSR, and SAM made substantial contribution to the conception of the work and the execution of the analysis; LVF, SK, DS, SAM, RL-R, JEC, AC-K, APS-S, MSR, ML made substantial contributions to the acquisition of the data, interpretation of the data, and revision of the work. Moreover, all authors have approved the final version and agree both to be personally accountable for the author’s own contributions and to ensure that questions related to the accuracy or integrity of any part of the work, even ones in which the author was not personally involved, are appropriately investigated, resolved, and the resolution documented in the literature.

## Acknowledgements

We would like to acknowledge the participants in the Mexican Teachers Cohort study,

